# Correlation of Polymorphism CYP2R1 and CYP27B1 with 25(OH)D3 concentration in Jakarta Healthy Population

**DOI:** 10.1101/2025.04.06.25325318

**Authors:** Margareta Amelia, Isadora Gracia, Linawati Hananta

## Abstract

**Background:** The correlation between vitamin D and the immune system began with the discovery of the VDR and CYP27B1 genes in almost all immune cells including T cells, B cells, neutrophils, macrophages and dendritic cells. The CYP2R1 rs10741657 and CYP27B1 rs10877012 genes have a quite important role in the vitamin D metabolism process starting from pre-vitamin D3 to 25(OH)D3 and then metabolized again to the active form 1,25(OH)2D3. Disruption of this cytochrome enzyme can reduce the concentration of 25(OH)D3.

**Objective:** Research that examines the correlation between biomolecular aspects of the CYP2R1 and CYP27B1 enzyme coding genes with blood serum 25(OH)D3 concentrations in a healthy population in Jakarta.

**Method:** This research uses an analytical observational study design with a cross-sectional study method. The independent variables are the CYP2R1 rs10741657 and CYP27B1 rs10877012 genes. DNA genotyping from blood using the polymerase chain reaction - restriction fragment length polymorphism (PCR-RFLP) method on 95 samples. DNA visualization using electrophoresis and gel-doc techniques. The dependent variable is the concentration of 25(OH)D3 which was measured using the Enzyme-Linked Immunosorbent Assay (ELISA) method. Statistical analysis was carried out using SPSS 22.

**Results:** The mean concentration of 25(OH)D3 for the group with the mutant genotype (AA) was 19.15 ng/mL + 19.61 indicating lower levels compared to the non-mutant genotype (GA/GG) with a mean concentration of 25(OH).)D3 32.41 ng/mL + 30.33 and 21.21 ng/mL + 18.39. The CYP27B1 rs10877012 mutant (GG) genotype had a mean 25(OH)D3 concentration of 23.58 ng/mL + 16.36, which was also lower than the 25(OH)D3 concentration of the non-mutant group (GT/TT) with a mean concentration value of 25(OH)D3 for the GT genotype 26.67 ng/mL + 20.87 and the TT genotype with a concentration of 29.27 ng/mL + 32.45. Analysis of the correlation between the CYP2R1 rs1074165 gene polymorphism and 25(OH)D3 concentration p value = 0.022, while the CYP27B1 rs10877012 gene polymorphism with 25(OH)D3 concentration obtained p value = 0.735.

**Conclusion:** There is a significant correlation between the CYP2R1 rs10741657 gene polymorphism and 25(OH)D3 concentration, while the CYP27B1 rs10877012 gene polymorphism does not have a significant correlation with 25(OH)D3 concentration.

## Introduction

Vitamin D is an important micronutrient that regulates the function of the body’s immune cells. This vitamin is reported to be able to stimulate the innate immune response by increasing the chemotaxis process, macrophage phagocytic response, and the production of antimicrobial proteins such as cathelicidin. When the body experiences vitamin D deficiency, it is more susceptible to dysregulation of the immune system and is more at risk of experiencing a cytokine storm.^1-5^ Vitamin D has an important role in modulating the immune system. This theory is supported by research by Ferreira *et al*. (2020) and Gaudet *et al*. (2022) that vitamin D has the ability to stimulate the innate immune response by increasing the chemotaxis process, phagocytic response from macrophages, and the production of antimicrobial proteins such as cathelicidin.^6,7^ Vitamin D also suppresses adaptive immunity by inhibiting the maturation of dendritic cells and reducing the ability to produce antigens presented to CD4.^1,8-10^

The metabolism of vitamin D into its active form, namely 25(OH)D3, is influenced by various cytochrome (CYP) P450 enzymes.^9^ In humans there are 57 genes that express the cytochrome (CYP) P450 enzyme. Any mutation in this CYP gene can cause serious health problems, one of which is vitamin D deficiency. ^11,12^ The first stage of vitamin D metabolism starts with the enzymatic process of forming vitamin D3 in the skin. The vitamin D3 that is formed will bind to vitamin D-binding protein (VDBP) in the blood vessels and then be carried to the liver. Vitamin D3 will first undergo hydroxylation in the liver by enzymes produced by cytochrome P450 (CYP2R1, CYP3A4, CYP27A1 and CYP2J3), especially CYP2R1, namely the 25-hydroxylase enzyme which can convert vitamin D3 into 25-hydroxy cholecalciferol (25(OH)D3). 25(OH)D3 is the main form that circulates in the blood, then binds to VDBP and is transported to the kidneys. In the renal proximal tubule, 25(OH)D3 is hydroxylated at the 1α position by the enzyme 1α-hydroxylase which converts 25(OH)D3 to the active form calcitriol (1α,25(OH)_2_D3). The 1α-hydroxylase enzyme is produced by cytochrome P450, namely CYP27B1.^1,9,13^

Vitamin D metabolism is regulated by the *CYP2R1, CYP27B1, CYP24A1* and *VDR* genes. The promoter region in the CpG islands of this gene is susceptible to mutations triggered by DNA methylation. The occurrence of this mutation can cause a decrease in the concentration of 25(OH)D3 in the blood circulation.^25^ The *CYP2R1* gene in humans is located on chromosome 11 arm p region 15 band 2 (11p15.2) consisting of 5 exons and 6 introns and spanning approximately 15.5 kb. This gene includes 501 AA with a protein weight of 51 Kda. A transition mutation occurs in the exon 2 of the *CYP2R1* gene which cause a Pro → Leu substitution at amino acid 99 in the CYP2R1 protein and eliminates the activity of the vitamin D 25-hydroxylase enzyme.^24^ Martinez-Hernandez et al. (2021) have showed that the CYP2R1 rs10741657 polymorphism was associated with an increased risk of vitamin D insufficiency, the results of this study also stated that the CYP2R1 polymorphism was significantly associated with low 25(OH)D3 serum levels.^22^

The *CYP27B1* gene is involved in 1α-hydroxylation and it converts 25(OH)D3 to its active formin humans. The *CYP27B1* gene is located on chromosome 12 arm q region 14 band 1 (12q14.1) and consists of nine exons. The translation product is a protein containing 508 amino acids with an N-terminal mitochondrial signal sequence and a heme binding site. The most frequently chosen CYP27B1 SNP is rs10877012 because it is found in the promoter region and its location can influence transcription and translation processes.^17^ This study aims to analyze the correlation of *CYP2R1* rs10741657 and *CYP27B1* rs10877012 gene with 25(OH)D3 concentrations.

## Method

### Research Design

This is an analytical observational study with cross sectional design. The respondents involved 132 recipients of booster vaccination at the Vaccination Center of the Faculty of Medicine and Health Sciences (FKIK) UNIKA Atma Jaya, Jakarta. The blood samples were collected from the 132 participants who had signed an informed consent form. Sampling was carried out from March to April 2022, then the blood samples were stored at −20°C. The 25(OH)D3 concentration was determined using the ELISA method, and the genotyping of *CYP2R1* rs10741657 and *CYP27B1* rs10877012 was carried out using the PCR-RFLP method.

### Subject

Subject selection used the principle of non-probability sampling with consecutive sampling techniques. Inclusion criteria for the participants to be included in this study are: (1) being over 17 years old, (2) willing to become research subjects, and (3) signing the informed consent form. Participants with a history of consuming vitamin D for more than 12 months and history of liver and/or kidney disorders and other comorbid diseases were excluded.

### Statistic Analysis

SPSS 22 software was used to perform statistical analysis. Chi square test and Fisher exact test were performed to analyze the correlation between *CYP2R1* rs10741657 and *CYP27B1* rs10877012 gene polymorphisms with 25(OH)D3 concentrations. The P-value significance threshold is 0.05 (p < 0.05).

### Examination

The determination of gene polymorphism was carried out at the Integrated Laboratory of the Faculty of Medicine and Health Sciences (FKIK) Unika Atma Jaya. The centrifugation was performed on 132 collected blood samples and were subsequently stored at −20°C. The concentration of 25(OH)D3 was measured using the ELISA (Enzyme Linked Analysis Techniques Immunosorbent Assay) method (Elabscience).^14^ The results of the 25(OH)D3 concentration called optimal were >30 ng/ml, insufficiency was 21-30 ng/ml and deficiency was < 20 ng/ml.^16^ DNA isolation (the Wizard® Genomic DNA purification Kit [Promega Corporation, USA])^15^ had been carried out and followed by genotyping of CYP2R1 rs10741657 and CYP27B1 rs10877012 using the Polymerase Chain Reaction-Restriction Fragment Length Polymorphism (PCR-RFLP) method.

The CYP2R1 rs10741657 genotyping examination. The first stage was examining the DNA isolation of the CYP2R1 rs10741657 gene by PCR using the forward primer 5’-GGGAAGAGCAATGACATGGA-3’ and reverse 5’-GCCCTGGAAGACTCATTTTG-3’. The PCR cycle protocol begins with an initial denaturation phase at 94 ºC for 5 minutes, then the denaturation phase at 94 ºC for 30 seconds (35 cycles), an annealing phase at 57 ºC for 30 seconds, an extension phase at 72 ºC for 40 seconds, and ends with a final extension phase at a temperature of 72 ºC for 7 minutes.^16^ The examination of CYP27B1 rs10877012 genotyping, using a PCR process mixes the DNA isolation with the forward primer 5’-GCCTGTAGTGCCTTGAGAGG-3’ and reverse 5’-CAGTGGGGAATGAGGGAGTA3’. The CYP27B1 gene PCR cycle protocol begins with an initial denaturation phase at 95 ºC for 5 minutes, then the denaturation phase at 95 ºC for 30 seconds (35 cycles), an annealing phase at 60 ºC for 30 seconds, an extension phase at 72 ºC for 60 seconds, and final extension phase at 72 ºC for 10 minutes.^17^

After the PCR process, the restriction length polymorphism (RFLP) process using the Mn1I restriction enzyme to genotype CYP2R1 rs10741657 is continued. Then, the electrophoresis process will display the genotype bands GG (151bp, 105bp, 32 bp), AA (256bp, 32bp), and heterozygous GA (256 bp, 151bp, 105bp, 32bp). Next, PCR RFLP for the CYP27B1 rs10877012 gene using the Hinfl restriction enzyme is carried out through electrophoresis process to obtain the GG (138bp, 49bp), TT (187bp), and heterozygous GT (187bp, 138bp, 49bp) genotype bands.^16-18^

### Review the Subject’s Ethics and Informed Consent

This research has received ethical approval based on the Declaration of Helsinki Guidelines. All subjects have been given an explanation regarding this research, understood and signed their agreement with the informed consent. All research procedures have received ethical approval from the research ethics commission of the Faculty of Medicine and Health Sciences, Atma Jaya Catholic University of Indonesia with number 23/09/KEP-FKIKUAJ/2023 on September 24^th^ 2023.

## Results

The total number of study participants was 132 with 8 blood samples whose 25(OH)D3 concentration could not be calculated so they were excluded in the next process, namely DNA isolation and determination of CYP2R1 rs10741657 and CYP27B1 rs10877012 gene polymorphisms. Among the 124 samples, only 95 samples had sufficient DNA concentration (>100ng/uL) for PCR - RFLP. (Figure 1.).

**Figure 1.**
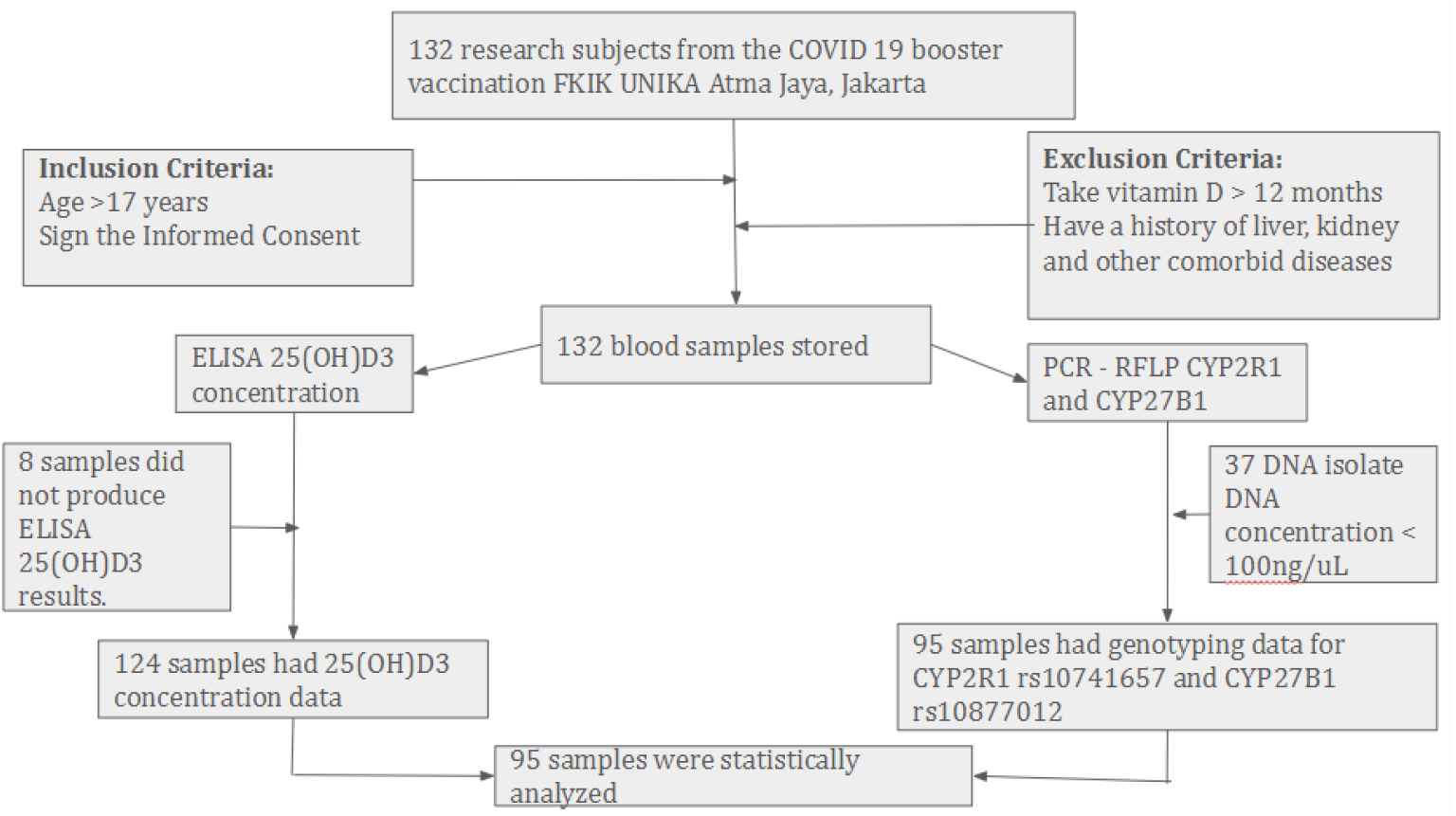
Research Flow

Tables 1 and 2 below show the characteristics of 95 subjects consisting of 43 people (45.3%) men and 52 people (54.7%) women, ranging from 18 to 79 years old. The average body mass index (BMI) was 24.75 + 4.21 with the lowest value being 16.14 and the highest being 34.40. The range of abdominal circumference ranges from 60 cm to 113.20 cm with an average of 88.95 + 11.26.

**Table 1.** Demographic Profile, ADEK Vitamin Intake and Sun Exposure.

| Parameter | n | % |
| --- | --- | --- |
| **Gender** |  |  |
| Male | 43 | 45.3 |
| Female | 52 | 54.7 |
| **ADEK Vitamin Intake** |  |  |
| Yes | 43 | 45.3 |
| No | 52 | 54.7 |
| **Sun Exposure** |  |  |
| Yes | 56 | 58.9 |
| No | 39 | 41.1 |

**Table 2.** Profile of Measurement and Laboratory Examination Results.

| Parameter | Minimum | Maximum | Mean | Std. Deviation |
| --- | --- | --- | --- | --- |
| Age | 18.00 | 79.00 | 45.32 ± | 15.97 |
| Body Mass Index (BMI) | 16.14 | 34.40 | 24.75 ± | 4.21 |
| Abdominal Circumference | 60.00 | 113.20 | 88.95 ± | 11.26 |
| Cholesterol Total | 105.00 | 393.00 | 192.74 ± | 49.57 |
| High-density lipoprotein (HDL) | 31.00 | 100.00 | 50.13 ± | 11.99 |
| Low-density lipoproteins (LDL) | 38.00 | 280.00 | 116.70 ± | 42.31 |
| Triglyceride | 45.00 | 379.00 | 130.44 ± | 63.26 |
| 25(OH)D3 concentration | 5.15 | 164.50 | 27.54 ± | 26.88 |

As many as 43 people (45.3%) consumed vitamins A, D, E and K for less than 12 months. In regards to the sun exposure, 56 people (58.9%) stated that they did outdoor activities and were exposed to sunlight quite often, 39 people (41.1%) said otherwise and did less outdoor activities and got less sunlight exposure. (Table 1.)

Laboratory examination was also carried out on the participants of this study, and secondary data was obtained with a mean total cholesterol concentration of 192.74 + 49.57 mg/dl with a minimum value of 105 mg/dl and of 393 mg/dl maximum. Meanwhile, for HDL examination, the mean was 50.13 + 11.99 mg/dl, LDL with a mean of 116.70 + 42.31 mg/dl and triglycerides with a mean of 130.44 + 63.26. The examination of the 25(OH)D3 concentration using the ELISA method resulted in the lowest concentration value being 5.15 ng/mL and the highest being 164.50 ng/mL with an average 25(OH)D3 concentration of 27.54 + 26.88 ng/mL. (Table 2)

Blood samples obtained from 25(OH)D3 concentrations were followed by DNA isolation and PCR-RFLP. PCR-RFLP examination for the CYP2R1 rs10741657 gene resulted in band visualization for each polymorphism as shown in Figure 2. For the AA genotype, band visualization was obtained at 256 bp and 32 bp. For the GA genotype, the band visualization was at 256 bp, 151 bp., 105 bp, and 32 bp and GG genotype with band visualization at 151 bp, 105 bp and 32 bp.

**Figure 2.**
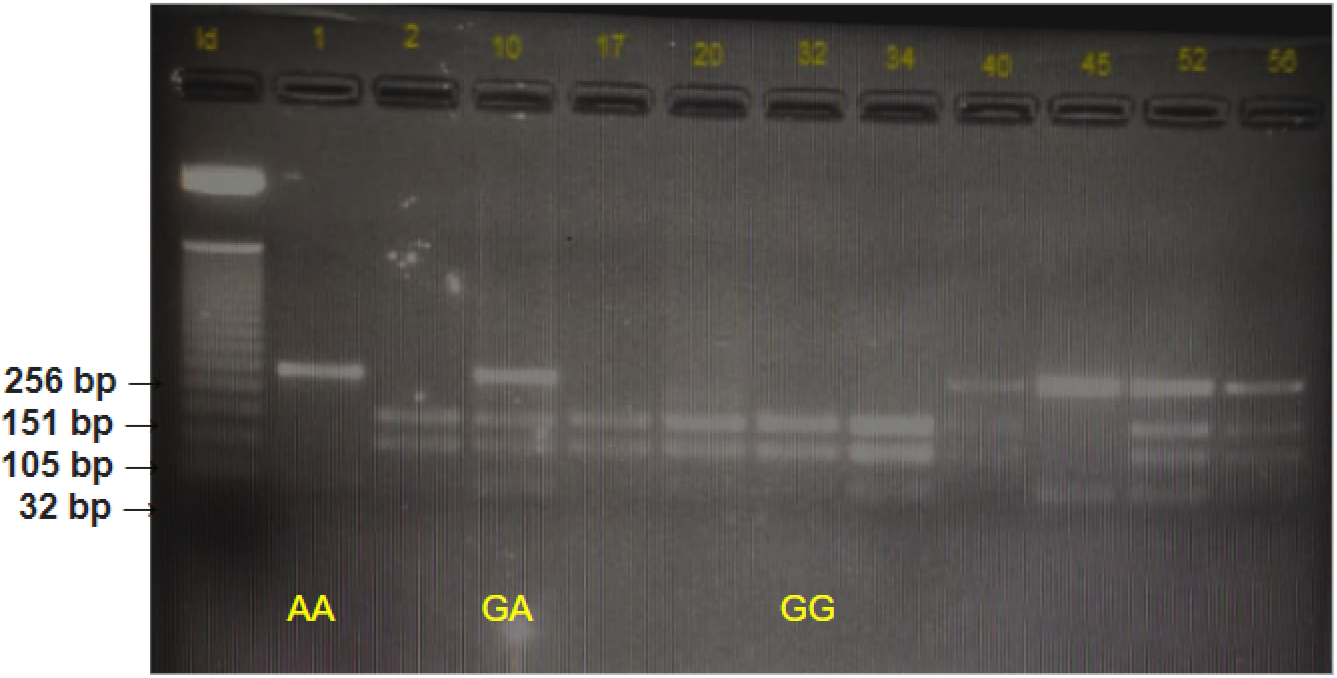
RFLP results of CYP2R1 rs10741657

The results of RFLP PCR visualization for the CYP27B1 rs10877012 gene polymorphism are shown in Figure 3. The GG genotype has band visualization at 138 bp and 49 bp, the GT genotype at 187 bp, 138 bp and 49 bp, and the TT genotype only at 187 bp.

**Figure 3.**
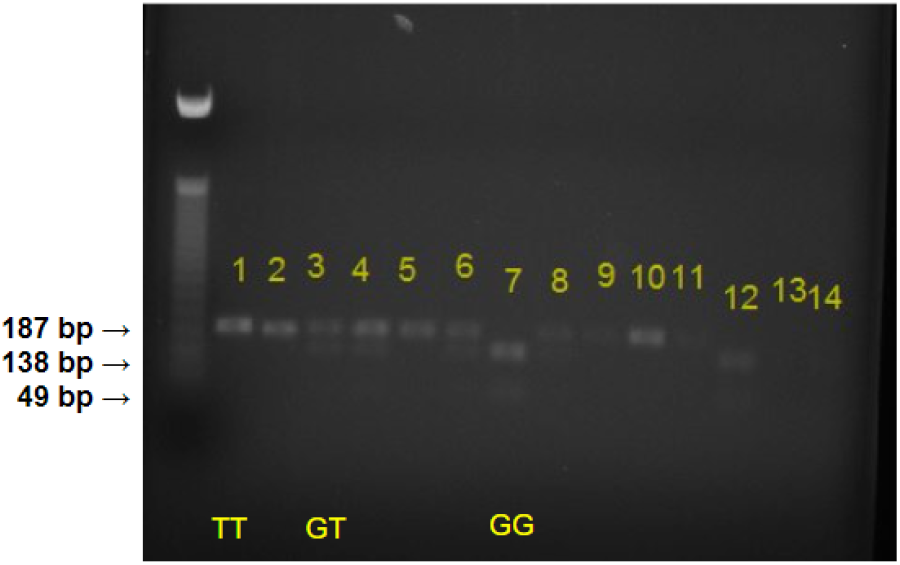
RFLP results of CYP27B1 rs10877012

Out of the 95 research subjects, 18 people (18.9%) had the mutant genotype (AA), 57 people (60.0%) heterogeneous (GA) and 20 people (21.1%) had the GG genotype for the CYP2R1 rs10741657 gene polymorphism. The population in this study complied with Hardy-Weinberg equilibrium (p=0.148). The mean 25(OH)D3 concentration for the group with the mutant genotype was 19.15 + 19.61, indicating lower levels compared to the non-mutant genotype (GA/GG) with 25(OH)D3 concentration mean values of 32.41 + 30.33 and 21.21 + 18.39. (Table 3.)

**Table 3.**
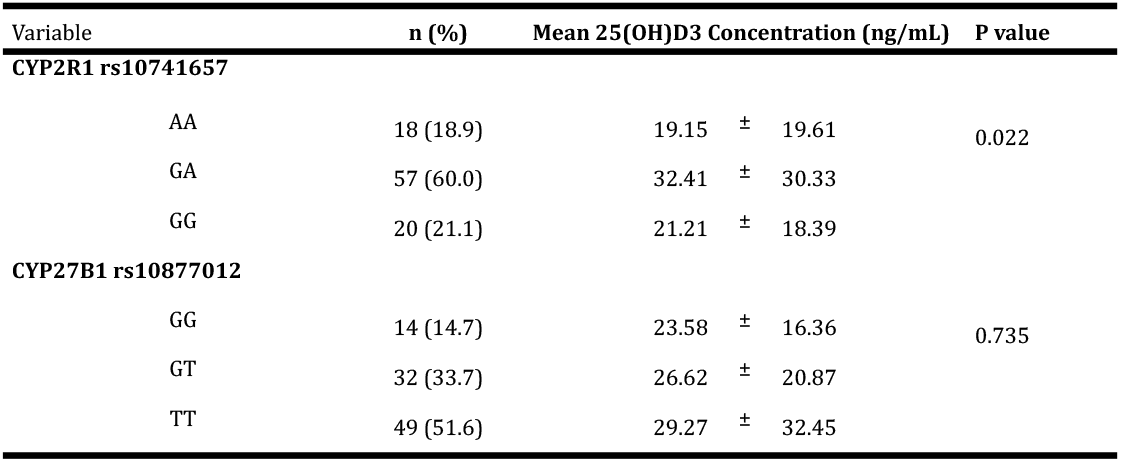
Frequency and Distribution of Gene Polymorphisms with 25(OH)D3 Concentration.

| Variable | n (%) | Mean 25(OH)D3 Concentration (ng/mL) |  | P value |
| --- | --- | --- | --- | --- |
| CYP2R1 rs10741657 |  |  |  |  |
| AA | 18 (18.9) | 19.15 | ± 19.61 | 0.022 |
| GA | 57 (60.0) | 32.41 | ± 30.33 |  |
| GG | 20 (21.1) | 21.21 | ± 18.39 |  |
| CYP27B1 rs10877012 |  |  |  |  |
| GG | 14 (14.7) | 23.58 | ± 16.36 | 0.735 |
| GT | 32 (33.7) | 26.62 | ± 20.87 |  |
| TT | 49 (51.6) | 29.27 | ± 32.45 |  |

The frequency distribution of the CYP27B1 rs10877012 gene polymorphism that have the mutant genotype (GG) was 14 people (14.7%) with an average 25(OH)D3 concentration of 23.58 + 16.36, which was also lower than the 25(OH)D3 concentration of the non-mutant group (GT /TT) with a mean value of 25(OH)D3 concentration for the GT genotype (32 people/33.7%) 26.67 + 20.87 and the TT genotype (49 people/51.6%) with a concentration of 29.27 + 32.45. (Table 3.) For the CYP27B1 rs10877012 gene. This study population also complies with Hardy-Weinberg equilibrium (p=0.099).

The correlation between the CYP2R1 rs10741657 genotype polymorphism has a significant relation to 25(OH)D3 concentration with p value = 0.022, while the CYP27B1 rs10877012 genotype polymorphism does not have a significant correlation with 25(OH)D3 concentration because p value = 0.735 (p > 0.05). (Table 3)

The results of statistical analysis showed that vitamin D deficiency in the group of participants with the CYP2R1 rs1074157 mutant (AA) genotype was 1.48 times higher than the GA/GG genotype (OR 1.484; 95% CI 0.530 - 4.157). Meanwhile, the CYP27B1 rs10877012 mutant (GG) genotype also showed 1.45 times higher levels of vitamin D deficiency compared to the GT/TT genotype (OR 1.455; 95% CI 0.466 - 4.537). (Table 4.)

**Table 4.** Correlation of CYP2R1 rs10741657 and CYP27B1 rs10877012 Genotype Polymorphism with 25(OH)D3 concentration.

|  | 25(OH)D3 concentration |  | OR | CI 95% |  | P value |
| --- | --- | --- | --- | --- | --- | --- |
|  | <30 ng/mL | >30 ng/mL |  | Lower | Upper |  |
| GENOTYPE | n (%) | n (%) |  |  |  |  |
| **CYP2R1 rs10741657** |  |  | 1.484 | 0.530 | 4.157 | 0.311 |
| GG / GA (non mutant) | 31 (32.6) | 46 (48.4) |  |  |  |  |
| AA (mutant) | 9 (9.5) | 9 (9.5) |  |  |  |  |
| **CYP27B1 rs10877012** |  |  | 1.455 | 0.466 | 4.537 | 0.358 |
| TT / GT (non mutant) | 33 (34.7) | 48 (50.5) |  |  |  |  |
| GG (mutant) | 7 (7.4) | 7 (7.4) |  |  |  |  |

## Discussion

Beside bone development, vitamin D is also closely related to the immune system. Studies on the correlation between vitamin D and the immune system began when there were reports of the presence of the VDR and CYP27B1 genes in almost all immune cells including T cells, B cells, neutrophils, macrophages and dendritic cells.^19^ The process of 25-hydroxylation vitamin D3 and vitamin D2 to become 25 (OH)D3 is assisted by the CYP2R1 gene. If there is an interference with this cytochrome, it can reduce the concentration of 25(OH)D3. Our research showed that more than 50% of total samples had optimal 25(OH)D3 concentrations (>30 ng/mL). (Table 4) The mean value of 25(OH)D3 concentration from the 95 samples was 27.54 ng/mL ± 26.88 (SD). (Table 2)

The condition of vitamin D deficiency in the blood in this study population is influenced by several factors including total cholesterol concentration and the CYP2R1 rs10741657 gene polymorphism, whereas several other factors such as sun exposure, the amount and type of food consumed as well as genetic variations found in other genes that are involved in the metabolism and catabolism of vitamin D as studied in this study, namely CYP27B1 rs10877012, does not have any significant effect. In contrast to the report by Mohhamed et al (2022), there was a relation between the CYP27B1 rs10877012 gene and that of the GT genotype. 25(OH)D3 concentrations were obtained which were much lower compared to the GG genotype (P=0.02) in the T2DM patient group. In addition, this study also revealed that significant higher body mass index (BMI), HbA1c levels, and fasting blood sugar concentrations were detected in individuals with vitamin D deficiency (P=0.009, P=0.001, and P=0.01) compared to those who have adequate vitamin D levels. From this study, the results showed that the CYP27B1 rs10877012 polymorphism is not a risk factor for T2DM but affects 25(OH)D3 concentrations in patients with diabetes and vitamin D deficiency conditions (P < 0.001).^20^

Studies on the correlation between 25(OH)D concentrations and CYP2R1 rs10741657 and CYP27B1 rs10877012 gene polymorphisms in healthy populations are still limited. One study was conducted by Misra et al (2023) involving 61 healthy respondents aged 18 to 45 years. This study proves that serum 25(OH)D3 concentration is significantly related to age and not to the presence of the CYP2R1 gene.^21^ Martinez et al (2021) studied the relation of 25(OH)D concentration, CYP2R1 rs10766197 polymorphism and CYP27B1 rs10877012 polymorphism between a group of Multiple Sclerosis (MS) patients compared to a control group. The results of this study showed that there were almost significant differences in the concentration of 25(OH)D in the CYP2R1 rs10766197 genotypes GG, GA, and AA (p = 0.08). For the TT genotype of the CYP27B1 rs10877012 polymorphism between MS patients and the control group, there was no significant difference between the CYP27B1 rs10877012 gene polymorphism and 25(OH)D3 concentration.^22^ In contrast to the results of research on the CYP2R1 rs10741657 and CYP27B1 rs10877012 polymorphisms in Indonesia (Meirina et al, 2023) in a population of pregnant women with latent tuberculosis infection showed that there was no significance between these two genes and the concentration of 25(OH)D3. (p = 0.541 and p = 0.057).^18^ Most studies related to the CYP2R1 rs10741657 and CYP27B1 rs10877012 polymorphisms were carried out in populations with various clinical disorders such as MS, T2DM, and pregnant women with latent tuberculosis infection. All these studies show various results. The causes of these various results include many factors that influence the conditions for the emergence of various diseases. Although almost all research attempts link to the presence of genetic variations in the vitamin D metabolism gene, it is thought that variations in this gene can also play a role in the risk of developing chronic infections, even malignancies apart from being related to 25(OH)D3 concentrations.

Our study showed that there was a correlation between the CYP2R1 rs10741657 genotype polymorphism and 25(OH)D3 concentration (p value = 0.022), while the CYP27B1 rs10877012 genotype polymorphism had no relation with 25(OH)D3 concentration because p value = 0.735 (p > 0.05). (Table 3) The CYP2R1 gene expresses the 25-hydroxylase enzyme playing a role in converting vitamin D3 into 25(OH)D3 in the liver. Meanwhile, the CYP27B1 gene codes for 1α-hydroxylation, which converts 25(OH)D3 into its active form 1,25(OH)2D3. CYP2R1 gene polymorphism causes a decrease in the effectiveness of the 25-hydroxylase enzyme so that the amount of vitamin D3 converted into 25(OH)D3 is reduced. The second metabolic process is the bioactivation of vitamin D3 which occurs in the kidneys. This process is assisted by the vitamin D 1α-hydroxylase enzyme (CYP27B1) which will hydrolyze 25(OH)D3 at the C1 position of the A chain to become 1,25-(OH)2D3. Regulation in the kidneys is very tight, and is carried out by parathyroid hormone. A decrease in calcium levels or an increase in phosphate levels will immediately trigger the production of parathyroid hormone by the parathyroid glands which increases the production of 1,25-(OH)2D3. This advanced metabolic process shows that the role of the CYP27B1 gene is not very significant with 25(OH)D3 concentrations.^1^

A total of 57 people (60.0%) of the 95 research subjects from the FKIK UAJ vaccine acceptor group had the GA genotype for the CYP2R1 rs10741657 gene polymorphism, 20 people had the GG genotype (21.10%), and the least number had the AA genotype (18 people) (18.9%). The low frequency of the AA (mutant gene) genotype compared to the GA/GG (non mutant gene) genotype in this study is in line with research by Meirina, et al (2023) and Harishankar et al (2021).^18,23^ Although there are similarities in the distribution of genotype frequencies, there are differences in the results of further analysis of the correlation between 25(OH)D concentration and genotype. This study showed that the highest mean concentration was found in the GA genotype (32.41 + 30.33) while the highest 25(OH)D3 concentration in Meirina et al’s report was detected in the AA genotype (58.90 + 59.19).^18^ Harishankar et al’s research showed the concentration of 25(OH)D3 significantly higher in the GA and AA genotype groups compared to the GG genotype. (GG vs GA; p=0.026; GG vs AA: HCs, p=0.036).^23^ This difference is due to the fact that in Meirina et al’s research, 1000 UI vitamin D supplementation was carried out for one month.^18^

Our study found that the CYP2R1 rs10741657 G allele was 97 (51.05%) while for the A allele it was found at 93 (48.95%). In accordance with the research by Lafi et al. (2015), the number of G alleles is greater than the A allele. The number of G alleles in the research of Lafi et al. (2015) 65.3% and the A allele was 34.7% of the total population of 762 participants. ^16^ Calculations of alleles and genotypes for the CYP2R1 rs10741657 and CYP27B1 rs10877012 genes are in accordance with the Hardy-Weinberg principle. This shows that within a population, the genetic variations that occur will remain constant from generation to generation without any other evolutionary influences.

The description of the CYP27B1 rs10877012 gene polymorphism in our study is that the TT genotype was the largest, which was found in 49 people (51.60%), followed by the GT genotype (32 people, 33.70%) and the GG genotype (14 people, 14.7%), with the highest mean 25(OH)D3 concentration. in the TT genotype: 29.27 ng/mL + 32.45 (SD). In contrast to the research conducted by Meirina, et al (2023), results showed that the highest frequency of the GT genotype was 46 people, followed by the GG genotype with 21 people, and the TT genotype with 17 people, with the highest 25(OH)D3 concentration in the GT genotype: 52.87 ng/mL + 47.31.^18^ Latacz et al, (2020)’s research on a caucasian population of colorectal cancer patients compared to a healthy population. In the healthy population the GT genotype was found to be the highest in 96 people (44%), then the GG genotype was 69 people (32%) and the TT genotype was 54 people (25%), with a frequency of the G allele of 53% and T of 47%.^17^ In our study, the frequency of the T allele of CYP27B1 rs10877012 was higher,:68.42%, compared to the G allele of 31.57%. This difference in allele frequency distribution is caused by different research subject populations. The research of Latacz et al. was conducted on a Caucasian population, while our research and that of Meirina et al., was conducted on an Indonesian population.

## Conclusion

This study proves that 25(OH)D3 concentrations are significantly associated with the *CYP2R1* rs10741657 gene polymorphism but not with the *CYP27B1* rs10877012 gene polymorphism. To explore the role of the *CYP27B1* gene in vitamin D3 metabolism, further research can be carried out linking the correlation between the *CYP27B1* gene and 1,25(OH)D3 concentrations.

## Data Availability

All data produced in the present study are available upon reasonable request to the authors

